# Clinical characteristics of imported and second-generation COVID-19 cases outside Wuhan, China: A multicenter retrospective study

**DOI:** 10.1101/2020.04.19.20071472

**Authors:** Puyu Shi, Guoxia Ren, Jun Yang, Zhiqiang Li, Shujiao Deng, Miao Li, Shasha Wang, Xiaofeng Xu, Fuping Chen, Yuanjun Li, Chunyan Li, Xiaohua Yang, Zhaofeng xie, Zhengxia Wu, Mingwei chen

## Abstract

**Background:** The mortality of COVID-19 differs between countries and regions. By now, reports on COVID-19 are largely focused on first-generation cases. This study aimed to clarify the clinical characteristics of imported and second-generation cases.

**Methods:** This retrospective, multicenter cohort study included 134 confirmed COVID-19 cases from 9 cities outside Wuhan. Epidemiological, clinical and outcome data were extracted from medical records and were compared between severe and non-severe cases. We further profiled the dynamic laboratory findings of some patients.

**Results:** 34.3% of the 134 patients were severe cases, and 11.2% had complications. As of March 7, 2020, 91.8% patients were discharged and one patient (0.7%) died. The median age was 46 years. The median interval from symptom onset to hospital admission was 4.5 (IQR 3-7) days. The median lymphocyte count was 1.1×10^9^/L. Age, lymphocyte count, CRP, ESR, DBIL, LDH, HBDH showed difference between severe and no-severe cases (all P<0.05). Baseline lymphocyte count was higher in the survived patients than in the non-survivor case, and it increased as the condition improved, but declined sharply when death occurred. The IL-6 level displayed a downtrend in survivors, but rose very high in the death case. Pulmonary fibrosis was found on later chest CT images in 51.5% of the pneumonia cases.

**Conclusion:** Imported and second-generation cases outside Wuhan had a better prognosis than initial cases in Wuhan. Lymphocyte count and IL-6 level could be used for evaluating prognosis. Pulmonary fibrosis as the sequelae of COVID-19 should be taken into account.

**Summary:** Imported and second-generation cases manifested less complications, lower fatality, and higher discharge rate than initial cases, which may be related to the shorter interval from symptom onset to hospital admission, younger age, and higher lymphocyte count of the imported and second-generation patients. Lymphocyte count and IL-6 level could be used as indicators for evaluating prognosis. Pulmonary fibrosis was found in later chest CT images in more than half of the pneumonia cases and should be taken into account.

## Introduction

In December 2019, a cluster of pneumonia cases of unknown cause occurred in the city Wuhan in China.[1] In early January, 2020, a novel betacoronavirus was isolated[2] from the bronchoalveolar lavage samples of the infected patients, and the pathogen was named severe acute respiratory syndrome coronavirus 2 (SARS-CoV-2; previously known as 2019 novel coronavirus, 2019-nCoV). In February 2020, WHO officially designated the syndrome as coronavirus disease 2019 (COVID-19).

Due to human-to-human transmission,[3, 4] COVID-19 has spread rapidly. As of March 27, 2020, a total of 82078 cases have been confirmed in China and 509164 cases have been reported in more than 200 countries and 5 continents.[5] The clinical spectrum of COVID-19 appears to be wide, including asymptomatic infection, mild respiratory tract illness, and severe pneumonia with respiratory failure and even death.[6] The mortality of COVID-19 is different among countries and regions, for instance 4.02% in China, 0% in Vietnam, 10.14% in Italy, 1.45% in USA,and 0.44% in Austrilia.[5]

So far, studies on the epidemic and clinical characteristics of COVID-19 have mainly concentrated on initial or first-generation cases. Information about imported and second-generation cases is limited. In this study, we focused on Shaanxi province as a region with imported and second-generation cases and described the clinical and laboratory characteristics of 134 COVID-19 cases in this province with a hope to provide some insight into the prevention and treatment of the disease in China and elsewhere.

## Methods

### Study design and participants

This retrospective study included 134 confirmed cases of COVID-19 admitted and treated in 10 designated hospitals across 9 cities (Xi’an, Ankang, Baoji, Hanzhong, Weinan, Xianyang, Shangluo, Yan’an, Tongchuan) in Shaanxi province from January 23, 2020 to March 7, 2020 (Supplementary Material). SARS-CoV-2 infection was defined in accordance with Version 7.0 of the guideline issued by the National Health Commission of the People’s Republic of China.[7]

### Data collection

The epidemiological, demographic, clinical, laboratory, and radiologic characteristics as well as treatment and outcome data were collected from patients’ electronic medical records using a standardised case report form. Clinical outcomes were followed up until March 7, 2020. The data were reviewed by a trained team of physicians. If information was not clear, the research team contacted the doctor responsible for treating the patient for clarification. Because of the urgent need to collect data regarding this emerging pathogen, requirement for informed consent was waived.

### Laboratory confirmation

Laboratory confirmation of COVID-19 was performed immediately after admission and verified by the Shaanxi Provincial Center for Disease Control and Prevention (CDC). A confirmed COVID-19 case was defined as a positive result to high-throughput sequencing or real-time reverse-transcriptase polymerase chain reaction (RT-PCR) assay for nasal and pharyngeal swab samples or sputum specimens.[8]

### Diagnostic criteria

The date of disease onset was defined as the day when the symptom was noticed. Fever was defined as axillary temperature above 37.3°C. ARDS was defined in accordance with the Berlin definition.[9] Acute kidney injury was identified based on the Kidney Disease: Improving Global Outcomes definition.[10] Cardiac injury was determined when the serum levels of cardiac biomarkers (e.g., troponin I/T) were above the 99th percentile upper reference limit or new abnormalities detected in electrocardiography and echocardiography.[11] Ventilator-associated pneumonia was determined referring to the guidelines for treatment of hospital-acquired and ventilator-associated pneumonia.[12] Severity of COVID-19 was categorized into non-severe group (mild and moderate) and severe group (sever and critically ill) based on Version 7.0 of the guideline issued by the National Health Commission of the People’s Republic of China.[7]

### Statistical analysis

The cohort of patients was divided into severe and non-severe cases. Continuous and categorical variables were expressed as median (IQR) and n (%), respectively. The Mann-Whitney test was used for continuous variables and χ^2^ test or Fisher’s exact test (when the data were limited) was used for categorical variables to compare differences between severe and non-severe cases where appropriate. All statistical analyses were performed with SPSS software, version 23.0. A two-sided α of less than 0·05 was considered statistically significant.

## Result

### Epidemiological and clinical characteristics

This study recruited a total of 134 patients from 9 cities in Shaanxi province, who were confirmed as SARS-CoV-2 infection from January 23, 2020 to March 7, 2020. The median age of the patients was 46 years old (IQR 34-58), ranging from 4 to 89 years, and more than half (69, 51.5%) were female (Table 1). Altogether 88 (65.7%) cases were non-severe and 46 (34.3%) were severe, including two critically ill cases (1.5%) with one patient unable to survive (0.7%) (Table 4). The age of severe patients were significantly older than that of non-severe patients (median, 56 years vs. 41 years, P <0.05). Moreover, the proportion of patients aged 65 or older was higher (32.6% vs. 5.7%, P <0.05), and the proportion of patients aged 14-30 was lower (4.3% vs. 21.6%, P <0.05) in the group of severe patients than in non-severe patients (Table 1).

**Table 1.** **Demographics, baseline characteristics of patients infected with COVID-19**

|  | Total<br>(N = 134) | Sevre<br>(N = 46) | Non-severe<br>(N = 88) | P value |
| --- | --- | --- | --- | --- |
| **Age, Median (IQR), years** | 46(34-58) | 56(46-66) | 41(29-50) | 0.000* |
| Age groups, No. (%) |  |  |  |  |
| <14 | 5(3.7) | 1(2.2) | 4(4.5) | 0.660 |
| 14-30 | 21(15.7) | 2(4.3) | 19(21.6) | 0.009* |
| 31-64 | 88(65.7) | 28(60.9) | 60(68.2) | 0.001* |
| ≥65 | 20(14.9) | 15(32.6) | 5(5.7) | 0.000* |
| **Sex, No. (%)** |  |  |  |  |
| Female | 69(51.5) | 21(45.7) | 48(54.5) | 0.328 |
| Male | 65(48.5) | 25(54.3) | 40(45.5) |  |
| **Exposure history, No. (%)** |  |  |  |  |
| Exposure history in Wuhan | 59(44) | 15(32.6) | 44(50) | 0.054 |
| Exposure history to individuals from Wuhan | 40(29.9) | 14(30.4) | 26(29.5) | 0.915 |
| Exposure history to COVID-19 patients | 20(14.9) | 9(19.6) | 11(12.5) | 0.276 |
| no definite epidemiological history | 15(11.2) | 8(17.4) | 7(8.0) | 0.1 |
| medical staff | 3(2.2) | 0(0) | 3(3.4) | 0.515 |
| Familial cluster | 71(53) | 22(47.8) | 49(55.7) | 0.272 |
| **Coexisting conditions, No. (%)** |  |  |  |  |
| Smoking | 14(10.4) | 6(13) | 8(9.1) | 0.680 |
| Drinking | 12(9.0) | 4(8.7) | 8(9.1) | 1.000 |
| Hypertension | 20(14.9) | 10(21.7) | 10(11.4) | 0.110 |
| Diabetes | 9(6.7) | 4(8.7) | 5(5.7) | 0.765 |
| Cardiovascular disease | 6(4.5) | 4(8.7) | 2(2.3) | 0.205 |
| Cerebrovascular disease | 6(4.5) | 3(6.5) | 3(3.4) | 0.698 |
| Respiratory system diseases | 5(3.7) | 4(8.7) | 1(1.1) | 0.087 |
| Malignancy | 5(3.7) | 2(4.3) | 3(3.4) | 1.000 |
| Chronic liver disease | 5(3.7) | 0(0) | 5(5.7) | 0.243 |
| Pregnency | 1(0.7) | 0(0) | 1(1.1) | 1.000 |
| HIV infection | 1(0.7) | 0(0) | 1(1.1) | 1.000 |
| Chronic kidney disease | 0(0) | 0(0) | 0(0) |  |
| Use of glucocorticoid/immunosuppressive agents | 0(0) | 0(0) | 0(0) |  |
| **Duration in hospital, Median (IQR), days** | 17(14-20) | 18(13-21) | 17(14-20) | 0.466 |
| **Onset of symptom to, Median (IQR), days** |  |  |  |  |
| Hospital admission | 4.5(3-7) | 5(3-7) | 4(2-7) | 0.395 |
| Diagnosis | 5(3-9) | 6(3-9) | 5(3-8) | 0.239 |

**Abbreviation:** Coronavirus disease-19 (COVID-19); IQR, interquartile range;
P values indicate differences between severe and non-severe patients. P < 0.05 was considered statistically significant.

**Table 4.** **Complications, Treatment and outcomes of patients infected with COVID-19**

|  | Total<br>(N = 134) | Severe<br>(N= 46) | Non-severe<br>(N=88) | P value |
| --- | --- | --- | --- | --- |
| **Complications, No. (%)** | 15(11.2) | 13(28.3) | 2(2.3) | 0.000* |
| Arrhythmia | 4(3.0) | 3(6.5) | 1(1.1) | 0.228 |
| Acute respiratory distress syndrome | 3(2.2) | 3(6.5) | 0(0) | 0.071 |
| Acute kidney injury | 3(2.2) | 2(4.3) | 1(1.1) | 0.563 |
| Ventilator-associated pneumonia | 2(1.5) | 2(4.3) | 0(0) | 0.116 |
| Multiple organ dysfunction syndrome | 2(1.5) | 2(4.3) | 0(0) | 0.116 |
| Shock | 1(0.7) | 1(2.2) | 0(0) | 0.343 |
| **Treatments** |  |  |  |  |
| **Antiviral treatment, No. (%)** | 129(96.3) | 46(100.0) | 83(94.3) | 0.243 |
| Lopinavir/ritonavir | 87(64.9) | 30(65.2) | 57(64.8) | 0.959 |
| Interferon alpha inhalation | 68(50.7) | 23(50.0) | 45(51.1) | 0.901 |
| Arbidol | 57(42.5) | 23(50) | 34(38.6) | 0.206 |
| Ribavirin | 44(32.8) | 22(47.8) | 22(25.0) | 0.008* |
| Chloroquine | 3(2.2) | 3(6.5) | 0(0) | 0.071 |
| Duration of antiviral treatment, Median (IQR), days | 13.0<br>(8.0-17.0) | 14.0<br>(9.0-18.3) | 12.0<br>(7.0-16.0) | 0.112 |
| Onset of symptom to antiviral, Median (IQR), days | 6.0<br>(4.0-9.0) | 6.0<br>(3.8-10.0) | 6.0<br>(4.0-8.8) | 0.542 |
| **Antibiotics treatment, No. (%)** | 103(76.9) | 40(87.0) | 63(71.6) | 0.045* |
| Duration of antibiotics treatment,<br>Median (IQR), days | 10.0<br>(7.0-14.0) | 11.0<br>(7.0-15.5) | 10.0<br>(6.3-14.0) | 0.325 |
| **Antifungal treatment, No. (%)** | 2(1.5) | 1(2.2) | 1(1.1) | 1.000 |
| **Glucocorticoid therapy, No. (%)** | 41(30.6) | 29(63.0) | 12(13.6) | 0.000* |
| Duration of glucocorticoid therapy,<br>Median (IQR), days | 3<br>(2.0-5.5) | 5<br>(3-6) | 3<br>(1-3) | 0.012* |
| Onset of symptom to glucocorticoid therapy,<br>Median (IQR), days | 6<br>(5.0-10.3) | 6<br>(5-12) | 6<br>(5-8) | 0.427 |
| **Gamma globulin, No. (%)** | 13(9.7) | 9(19.6) | 4(4.5) | 0.013* |
| Duration of Gamma globulin therapy,<br>Median (IQR), days | 4.0<br>(3.0-7.0) | 4.0<br>(2.5-7.0) | 5.0<br>(3.5-16.3) | 0.351 |
| **Oxygen inhalation, No. (%)** | 91(67.9) | 43(93.5) | 48(54.4) | 0.000* |
| **Non-invasive ventilation, No. (%)** | 2(1.5) | 2(4.3) | 0(0) | 0.116 |
| **Invasive mechanical ventilation, No. (%)** | 1(0.7) | 1(2.2) | 0(0) | 0.343 |
| **Extracorporeal membrane oxygenation, No. (%)** | 1(0.7) | 1(2.2) | 0(0) | 0.343 |
| **Continuous renal replacement therapy, No. (%)** | 1(0.7) | 1(2.2) | 0(0) | 0.343 |
| **Clinical outcome, No. (%)** |  |  |  |  |
| Discharge | 123(91.8) | 39(84.8) | 84(95.5) | 0.071 |
| Remained in hospital | 9(6.7) | 7(15.2) | 2(2.3) | 0.013* |
| Died | 1(0.7) | 1(2.2) | 0(0) | 0.343 |

P values indicate differences between severe and non-severe patients. P < 0.05 was considered statistically significant.

None of the patients had a history of exposure to the Huanan Seafood Market or wild animals. The majority of the cases were community-infected and three cases were hospital-infected. Of these patients, 59 (44%) resided in Wuhan or had short trips to Wuhan before the onset of COVID-19; 40 (29.9%) had close contact with someone from Wuhan; 20 (14.9%), including 3(2.2%) medical staff, had exposure to COVID-19 patients; 15 (11.2%) had no definite epidemiological history; and 71(53%) patients got infected as familial clustering (Table 1).

Of the 134 patients, 58 (43.3%) had one or more coexisting medical conditions, the most common of which was hypertension (14.9%), followed by diabetes (6.7%), cardiovascular disease (4.5%) and cerebrovascular disease (4.5%)(Table 1). The most common symptoms at onset were cough (96, 71.6%), followed by fever (87, 64.9%)(Table 2). The incidence of chest stuffiness and dyspnea differed between severe and no-severe cases (Table 2, both P < 0.05). The median interval from onset of symptoms to first hospital admission was 4.5 (IQR 3-7) days, and that to positive result of nucleic acid detection was 5 (IQR 3-9) days. The median duration from hospital admission to discharge was 17 days (IQR 14-20) (Table 1).

**Table 2.** **Symptomatic and radiological characteristics of patients infected with COVID-19**

|  | Total<br>(N = 134) | Sevre<br>(N = 46) | Non-severe<br>(N = 88) | P value |
| --- | --- | --- | --- | --- |
| **Symptoms, No. (%)** |  |  |  |  |
| Cough | 96(71.6) | 39(84.8) | 57(64.8) | 0.015* |
| Fever | 87(64.9) | 34(73.9) | 53(60.2) | 0.115 |
| Sputum | 61(45.5) | 27(58.7) | 34(38.6) | 0.027* |
| Fatigue | 55(41) | 23(50) | 32(36.4) | 0.128 |
| Anorexia | 45(33.6) | 19(41.3) | 26(29.5) | 0.170 |
| Chest stuffiness | 32(23.9) | 19(41.3) | 13(14.8) | 0.001* |
| Sore throat | 21(15.7) | 9(19.6) | 12(13.6) | 0.370 |
| Nausea and vomiting | 14(10.4) | 6(13) | 8(9.1) | 0.680 |
| Diarrhea | 13(9.7) | 7(15.2) | 6(6.8) | 0.210 |
| Muscle ache | 13(9.7) | 7(15.2) | 6(6.8) | 0.210 |
| Dyspnea | 12(9.0) | 8(17.4) | 4(4.5) | 0.031* |
| Headache | 12(9.0) | 4(8.7) | 8(9.1) | 1.000 |
| Chest pain | 10(7.5) | 4(8.7) | 6(6.8) | 0.963 |
| Dizziness | 9(6.7) | 4(8.7) | 5(5.7) | 0.765 |
| Rhinorrhoea | 7(5.2) | 2(4.3) | 5(5.7) | 1.000 |
| Palpitation | 6(4.5) | 4(8.7) | 2(2.3) | 0.205 |
| Abdominal pain | 4(3.0) | 1(2.2) | 3(3.4) | 1.000 |
| arthralgia | 3(2.2) | 3(6.5) | 0(0) | 0.071 |
| Confusion | 2(1.5) | 2(4.3) | 0(0) | 0.116 |
| Conjunctivitis | 0(0) | 0(0) | 0(0) |  |
| **Signs** |  |  |  |  |
| Highest temperature, No. (%) |  |  |  |  |
| <37.3°C | 47(35.1) | 13(28.3) | 34(38.6) | 0.232 |
| 37.3-38°C | 41(30.6) | 9(19.6) | 32(36.4) | 0.045* |
| >38°C | 46(34.3) | 24(52.2) | 22(25) | 0.002* |
| Respiratory rate, No. (%) |  |  |  |  |
| 12-20 | 77(57.5) | 17(37.0) | 60(68.2) | 0.001* |
| >21 | 57(42.5) | 29(63) | 28(31.8) | 0.001* |
| Heart rate, No. (%) |  |  |  |  |
| ≤100 | 109(81.3) | 32(69.6) | 77(87.5) | 0.001* |
| >100 | 25(18.7) | 14(30.4) | 11(12.5) | 0.011* |
| Systolic pressure, Median (IQR), mmHg | 120(110-139) | 130(120-144) | 120(110-130) | 0.001* |

**Table 2. (continued)**
|  | Total<br>(N = 134) | Sevre<br>(N = 46) | Non-severe<br>(N = 88) | P value |
| --- | --- | --- | --- | --- |
| **Chest CT findings, No. (%)** |  |  |  |  |
| No abnormal density shadow | 8(6.0) | 0(0) | 8(9.1) | 0.085 |
| Unilateral mottling and ground-glass opacity | 26(19.4) | 5(10.9) | 21(23.9) | 0.071 |
| Bilateral mottling and ground-glass opacity | 100(74.6) | 41(89.1) | 59(67.0) | 0.005* |
| Multiple patchy shadowing | 26(19.4) | 10(21.7) | 16(18.2) | 0.621 |
| Pulmonary consolidation | 26(19.4) | 12(26.1) | 14(15.9) | 0.157 |
| Air bronchogram | 5(3.7) | 3(6.5) | 2(2.3) | 0.452 |
| White lung | 2(1.5) | 2(4.3) | 0(0) | 0.116 |
| Pleural effusion | 6(4.5) | 5(10.9) | 1(1.1) | 0.032* |
| Pneumothorax | 1(0.7) | 1(2.2) | 0(0) | 0.343 |
| Pulmonary fibrosis | 69(51.5) | 30(65.2) | 39(44.3) | 0.022* |

P values indicate differences between severe and non-severe patients. $P < 0.05$ was considered statistically significant.

Upon admission, measures of vital signs were recorded for all patients. The incidences of temperature >38°C, respiratory rate > 21 breaths per minute, heart rate > 100 beats per minute and median systolic pressure showed difference between severe and no-severe cases (Table 2, all P < 0.05). All of these are higher in severe cases compared to in non-severe cases.

### Radiological and laboratory findings

94.0% (126/134) of the patients showed abnormal chest CT images, consisting of 26 cases (26/134, 19.4%) of unilateral pneumonia and 100 cases (100/134, 74.6%) of bilateral pneumonia, with ground-glass opacity as the typical hallmark finding. Among the patients, 26 patients (19.4%) showed multiple patchy shadowing, 26 cases (19.4%) subsegmental consolidation with air bronchogram (5, 3.7%), with 2 cases (1.5%) having progressed to “white lung”. Additionally, pleural effusion occured in 6 patients (4.5%) and pneumothorax occurred in 1 patient (0.7%). When the shadow or consolidation were resolved, fibrous stripes or pulmonary fibrosis were found on later chest CT images of 69 (51.5%) patients (Table 2). Moreover, the incidences of bilateral pneumonia, pleural effusion and pulmonary fibrosis were higher in severe cases than in non-severe cases (Table 2, all P < 0.05).

Upon admission, 25 (18.7%) of the patients showed leucopenia (white blood cell count <3.5×10^9^/L) and 51 (38.1%) showed lymphopenia (lymphocyte count <1.1×10^9^ /L). In most patients, leukocytes (107, 79.9%) and lymphocytes (82, 61.2%) were within the normal ranges. Only two patients (1.5%) had increased leukocytes and one patient (0.7%) had elevated lymphocytes. The median values of C-reactive protein (10.0, IQR 9.0-38.3, mg/L), erythrocyte sedimentation rate (38.5, IQR 17.8-74.8, mm/h), interleukin-6 (8.1, IQR 5.0-23.0, pg/ml), and direct bilirubin (4.9, IQR 3.3-7.5, umol/L) elevated in the patients. The median partial pressure of oxygen level was 80 mmHg (IQR 67-92), and the median of oxygenation index (PaO_2_:FiO_2_) was 255 mmHg (IQR 210-307) (Table 3).

**Table 3.** **laboratory findings of patients infected with COVID-19. Values are medians (interquartile ranges) unless stated otherwise**

|  | Normal Range | Total (N = 134) | Severe (N = 46) | Non-severe (N = 88) | P value |
| --- | --- | --- | --- | --- | --- |
| **Blood routine** |  |  |  |  |  |
| White blood cell count ($\times 10^9/L$) | 3.5-9.5 | 4.7 (3.6-5.8) | 4.7 (3.8-5.8) | 4.7 (3.6-5.9) | 0.870 |
| <3.5, No. (%) |  | 25(18.7) | 7(15.2) | 18(20.5) | 0.460 |
| 3.5-9.5, No. (%) |  | 107(79.9) | 37(80.4) | 70(79.5) | 0.903 |
| >9.5, No. (%) |  | 2(1.5) | 2(4.3) | 0(0) | 0.116 |
| Lymphocyte count ($\times 10^9/L$) | 1.1-3.2 | 1.1 (0.9-1.4) | 1.0 (0.7-1.4) | 1.2 (0.9-1.5) | 0.031* |
| <1.1, No. (%) |  | 51(38.1) | 24(52.1) | 27(30.7) | 0.015* |
| 1.1-3.2, No. (%) |  | 82(61.2) | 22(47.8) | 60(68.2) | 0.022* |
| >3.2, No. (%) |  | 1(0.7) | 0(0) | 1(1.1) | 1.000 |
| Neutrophil count ($\times 10^9/L$) | 1.8-6.3 | 2.9 (2.2-4.1) | 3.0 (2.3-4.3) | 2.9 (2.1-4.0) | 0.583 |
| Platelet count count ($\times 10^9/L$) | 125-350 | 171 (132-219) | 182 (145-232) | 169 (128-213) | 0.183 |
| Haemoglobin (g/L) | 115-150 | 134 (121-148) | 139 (125-148) | 132 (121-149) | 0.485 |
| **Inflammatory mediator** |  |  |  |  |  |
| C-reactive protein (mg/L) | 0-10 | 10.0 (9.0-38.3) | 24.4 (10.0-67.4) | 9.0 (8.9-20.4) | 0.000* |
| Erythrocyte sedimentation rate(mm/h) | 0-20 | 38.5 (17.8-74.8) | 43.0 (26.0-84.0) | 30.0 (13.0-70.0) | 0.019* |
| Procalcitonin (ng/ml) | 0-0.5 | 0.06 (0.41-0.1) | 0.07 (0.05-0.11) | 0.05 (0.04-0.01) | 0.068 |
| Interleukin-6 (pg/ml) | 0-7 | 8.1 (5.0-23.0) | 23.0 (7.2-49.7) | 5.7 (5.0-9.9) | 0.063 |
| **Blood biochemistry** |  |  |  |  |  |
| Alanine aminotransferase (U/L) | 7-40 | 26 (16-52) | 25.4 (18.0-43.0) | 27.4 (13.3-53.8) | 0.847 |
| Aspartate aminotransferase (U/L) | 13-35 | 26 (20-41) | 27.0 (21.5-41.5) | 26.0 (19.0-40.8) | 0.235 |
| γ-glutamyl transpeptidase (U/L) | 7-45 | 26 (14-54) | 31.1 (18.5-55.0) | 22.7 (13.0-52.8) | 0.174 |
| Alkaline phosphatase (U/L) | 50-100 | 68 (57-87) | 68.0 (56.5-94.0) | 69.0 (57.1-85.8) | 0.898 |
| Total bilirubin (umol/L) | 0-17 | 14.7 (9.4-21.0) | 16.6 (10.8-20.4) | 14.2 (9.2-21.6) | 0.619 |
| Direct bilirubin (umol/L) | 0-3.4 | 4.9 (3.3-7.5) | 6.7 (3.9-8.4) | 4.4 (3.2-6.8) | 0.010* |
| Indirect bilirubin (umol/L) | 3.4-17.1 | 9.7 (6.2-14.6) | 9.8 (6.5-13.9) | 9.0 (6.2-15.2) | 0.745 |
| Albumin (g/L) | 40-55 | 40.1 (35.0-43.5) | 38.0 (33.2-42.5) | 41.2 (36.6-43.8) | 0.009* |
| Blood urea nitrogen (mmol/L) | 3.1-7.5 | 3.6 (3.0-4.5) | 3.8 (2.9-5.7) | 3.5 (3.0-4.3) | 0.099 |

**Table 3. (continued)**
|  | Normal Range | Total (N = 134) | Severe (N = 46) | Non-severe (N = 88) | P value |
| --- | --- | --- | --- | --- | --- |
| Creatinine (umol/L) | 41-111 | 60.0 (50.4-73.0) | 61.5 (52.8-76.0) | 58.0 (50.0-72.0) | 0.356 |
| Potassium (mmol/L) | 3.5-5.3 | 3.9 (3.7-4.3) | 3.8 (3.6-4.3) | 4.0 (3.7-4.4) | 0.081 |
| Sodium (mmol/L) | 137-147 | 139.3 (136.3-141.5) | 139.0 (135.0-140.3) | 139.5 (137.4-141.8) | 0.036* |
| Glucose (mmol/L) | 3.9-6.1 | 5.5 (4.9-6.3) | 6.0 (5.2-8.4) | 5.4 (4.7-6.2) | 0.001* |
| Creatine kinase (U/L) | 40-200 | 4 (2-6) | 3.5 (2.0-6.0) | 4.0 (2.0-6.0) | 0.452 |
| Creatine kinase-MB (U/L) | 0-24 | 8 (1-13) | 8.0 (0.8-12.8) | 8.0 (1.0-12.6) | 0.953 |
| Lactate dehydrogenase (U/L) | 120-250 | 206 (150-259) | 235.0 (204-300) | 177.0 (140.0-248.5) | 0.001* |
| Hydroxybutyrate dehydrogenase (U/L) | 72-182 | 153 (123-196) | 185 (149-237) | 136 (114-185) | 0.000* |
| Pro-brain natriuretic peptide (pg/mL) | 0-125 | 29.6 (14.2-84.6) | 39.5 (22.6-169.4) | 25.6 (12.6-54.6) | 0.034* |
| **Coagulation function** |  |  |  |  |  |
| D-dimer (mg/L) | 0-1.0 | 0.5 (0.3-0.7) | 0.5 (0.3-1.2) | 0.5 (0.3-0.7) | 0.338 |
| Prothrombin time (s) | 11-14 | 11.7 (10.7-13.5) | 11.7 (11.0-14.1) | 11.8 (10.7-13.4) | 0.725 |
| Activated partial thromboplastin time (s) | 28-43.5 | 31.6 (27.9-36.1) | 32.6 (27.7-38.2) | 31.2 (28.2-36.0) | 0.592 |
| Fibrinogen (g/L) | 2-4 | 3.3 (2.3-4.0) | 3.7 (2.3-4.3) | 3.2 (2.3-3.9) | 0.121 |
| **Blood Gas Analysis** |  |  |  |  |  |
| PH | 7.35-7.45 | 7.44 (7.41-7.47) | 7.46 (7.42-7.48) | 7.41 (7.40-7.44) | 0.000* |
| PaO<sub>2</sub> (mmHg) | 75-100 | 80 (67-92) | 70 (60-80) | 93 (88-119) | 0.000* |
| PaO<sub>2</sub>:FiO<sub>2</sub> (mmHg) |  | 255 (210-307) | 239 (199-273) | 433 (324-486) | 0.000* |
| PaCO<sub>2</sub> (mmHg) | 35-45 | 36 (33-39) | 35 (32-38) | 37 (33-40) | 0.152 |
| HCO<sub>3</sub><sup>-</sup> (mmol/L) | 22-26 | 24.2 (22.4-26.1) | 24.5 (22.2-27.1) | 23.5 (22.6-25.5) | 0.426 |
| **Co-infection, No. (%)** |  |  |  |  |  |
| Bacteria |  | 23(17.2) | 8(17.4) | 15(17.0) | 0.960 |
| Fungus |  | 3(2.2) | 2(4.3) | 1(1.1) | 0.563 |
| Other viruses |  | 6(4.5) | 0(0) | 6(6.8) | 0.170 |

**Abbreviation:** PaO<sub>2</sub>, partial pressure of oxygen; FiO<sub>2</sub>, fraction of inspired oxygen; PaCO<sub>2</sub>, partial pressure of carbon dioxide;
P values indicate differences between severe and non-severe patients. P < 0.05 was considered statistically significant.

A number of laboratory parameters showed higher values in severe patients as compared with in non-severe patients (Table 3), including C-reactive protein (CRP), erythrocyte sedimentation rate (ESR), direct bilirubin (DBIL), glucose, lactate dehydrogenase (LDH), hydroxybutyrate dehydrogenase (HBDH), and pro-brain natriuretic peptide (Table 3, all P < 0.05). In addition, lymphocyte count, albumin, PaO_2_, and PaO_2_:FiO_2_ were comparatively lower in severe patients than in non-severe patients (Table 3, all P < 0.05).

### Complications, treatment and outcomes

During hospitalization, 15 (11.2%) of the patients had complications, including arrhythmia (4, 3.0%), acute respiratory distress syndrome (3, 2.2%), acute kidney injury (3, 2.2%), ventilator-associated pneumonia (2, 1.5%), multiple organ dysfunction syndrome(2, 1.5%) and shock (1, 0.7%). Most of the complications (13 out of 15, 86.7%) occurred in the group of severe cases and the incidence of complications was comparatively higher in severe cases than in non-severe cases (28.3% vs. 2.3%, P< 0.05) (Table 4).

As for therapeutic management, 91 (67.9%) patients received oxygen inhalation, The two critical illness cases (1.5%) were treated with noninvasive ventilation (NIV), of whom one switched to invasive mechanical ventilation (IMV), extracorporeal membrane oxygenation (ECMO) and continuous renal replacement therapy (CRRT) as salvage therapy, and the other died before switching to invasive mechanical ventilation (Table 4).

Of the 134 patients, 23 (17.2%) experienced a secondary bacteria infection, 3(2.2%) were detected as positive for secondary fungus infection, and 6(4.5%) had other viruses infection (Table 3). Empirical single antibiotic treatment, mainly moxifloxacin, was given to 103 patients (76.9%), with a median duration of 10 days (IQR 7-14). Most patients (129, 96.3%) received antiviral therapy (median duration 13 days, IQR 8-17), including lopinavir/ritonavir (87, 64.9%), interferon alpha inhalation (68, 50.7%), arbidol (57,42.5%), ribaviron (44, 32.8%), and chloroquine (3, 2.2%). The median interval from onset of symptoms to antiviral therapy was 6.0 (IQR 4-9) days (Table 4). Additionally, 2 patients (1.5%) received antifungal treatment (Table 4).

Glucocorticoid therapy (median duration 3 days, IQR 2.0-5.5) was performed in 41 patients (30.6%), the duration of which is remarkably longer in severe cases than in non-severe cases (median 5 vs. 3, P<0.05). The median interval from onset of symptoms to glucocorticoid therapy was 6 days (IQR 5.0-10.3). In addition, 13 cases (9.7%) was supported with gamma globulin (median 4 days, IQR 3.0-7.0). Significantly more severe cases were given oxygen inhalation, antibiotics, systematic corticosteroid and gamma globulin (all P<0.05, Table 4).

By March 7, 2020, 123(91.8%) of the 134 patients had been discharged and one critical patient (0.7%) had died. The remaining 9 patients (6.7%) still under treatment were largely severe cases (7 out of 46 severe, 15.2% vs. 2 out of 88 non-severe, 2.3%, P<0.05) (Table 4). Fitness for discharge was based on abatement of fever for at least three days, significantly improved respiratory symptoms, and negative result for two consecutive respiratory pathogenic nucleic acid tests (sampling interval at least 1 day).

### Dynamic Profile of Laboratory Findings in Patients With COVID-19

To determine the major clinical features during COVID-19 progression, the dynamic changes of six clinical laboratory parameters, namely, lymphocyte, interleukin-6 (IL-6), CRP, LDH, HBDH, and DBIL, were monitored every other day from day 1 to day 8 after hospital admission. By January 28, 2020, data of the complete clinical course from seven patients, including five randomly selected discharged patients, the critical case managed with ECMO, and the non-survivor case, were analyzed (Figure 1). Baseline lymphocyte count was significantly higher in survivors than in the non-survivor patient. In survivors, the lymphocyte count was lowest on day 5 after admission and increased gradually during hospitalization. Whereas, the non-survivor patient developed more severe lymphopenia (0.19×10^9^/L) over time. The level of IL-6 in survivors displayed a gradual decrease to normal range with the condition improved, but increased unexpectedly to a very high value (5001pg/ml) before death in the non-survivor case.

**Figure 1:**
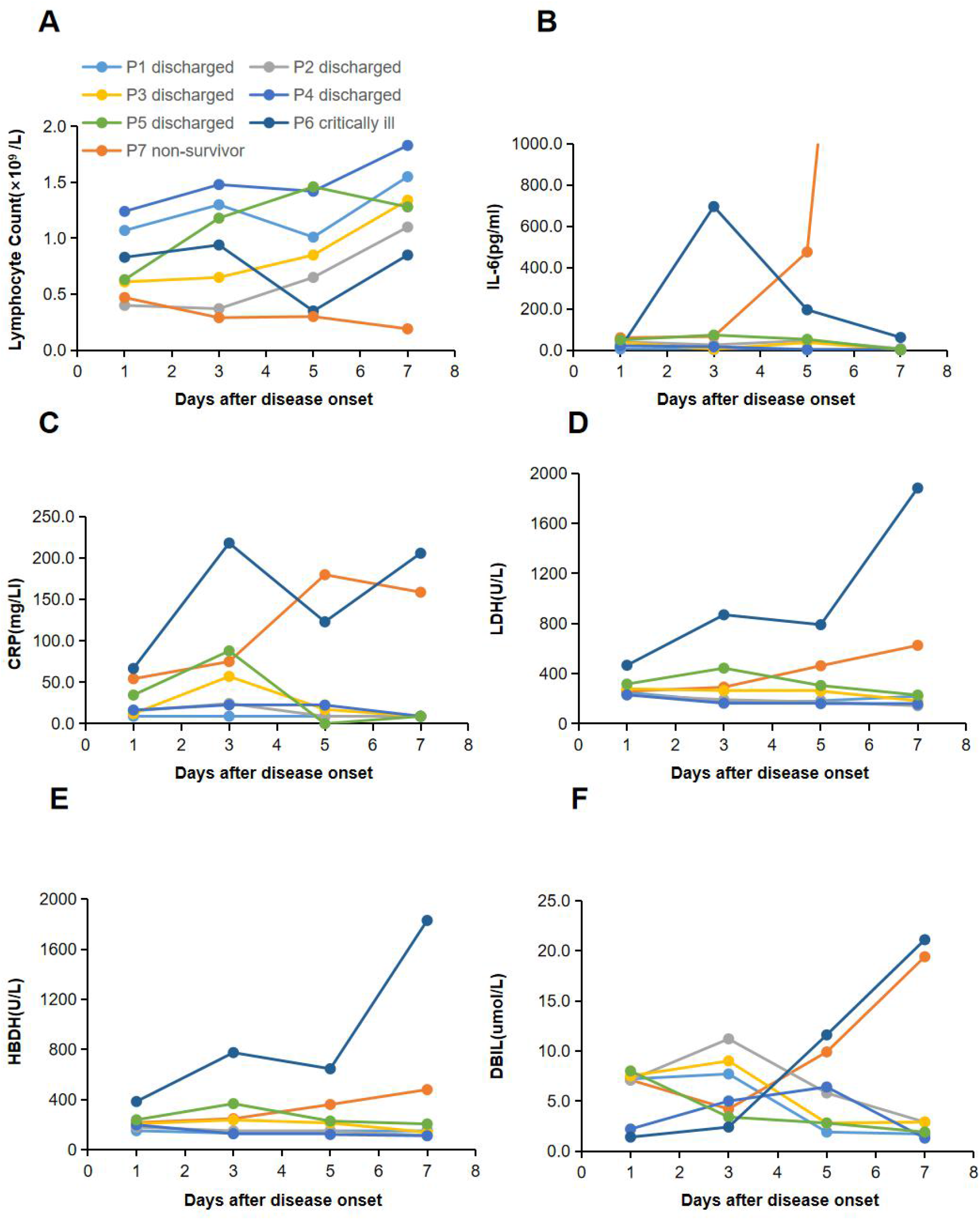
Dynamic Profile of Laboratory Findings in Patients With COVID-19. Timeline charts illustrated the dynamic changes of 6 laboratory markers (lymphocyte, IL-6, CRP, LDH, HBDH, DBIL) in 7 COVID-19 patients (5 discharged patients, 1 critical case managed with ECMO, 1 non-survivor) every other day from illness onset. (A-F) Dynamic changes of lymphocyte (A), IL-6 (B), CRP (C), LDH (D), HBDH (E), DBIL (F). The descriptive curve of individual patient: discharged/cured cases: P1, P2, P3, P4, P5; critically ill cases: P6; non-survivor: P7 was displayed. **Abbreviation:** IL-6: Interleukin-6; CRP: C-reactive protein; LDH: Lactate dehydrogenase; HBDH: Hydroxybutyrate dehydrogenase; DBIL: Direct bilirubin. COVID-19: Coronavirus disease-19.

Compared with those in the recovered patients, levels of CRP, LDH, HBDH, and DBIL in the two critically ill patients were higher throughout the clinical course (Figure 1). In the recovered patients,the levels of all the four markers reached the peak on day 3 after admission and decreased subsequently during recovery. In the two critically ill cases, the levels increased rapidly from day 3 with condition deterioration.

## Discussion

No patient enrolled in this multicenter study had been exposed to the Huanan Seafood Market or wild animals. The majority of the patients had a clear history of having been in Wuhan or in close contact with individuals from Wuhan or patients with COVID-19, and a large number of them were family clusters (53%). These data have provided further evidence that SARS-CoV-2 has a strong ability for human-to-human transmission.[3, 4] Besides, the fact that 11.2% of the patients had no definite epidemiological history might suggest the possibility of transmission from asymptomatic individuals. At present, confirmed COVID-19 patients are the main source of infection, but what deserves particular attention is asymptomatic transmission, which could be another source of infection. Moreover, 44% of the patients had a histroy of exposure in Wuhan, but 56% had never been to Wuhan and had been infected outside Wuhan. This suggests a gradual shift of imported infection to second-generation local infection.

The percentage of male patients in our data was 48.5%, different from the male patient predominence reported in two studies on Wuhan cases (73% in Huang et al[13] and 68% in Chen et al[14]). In this study, the male-female ratio was approximately 1:1.06, with no difference between severe and non-severe cases. This finding is contradicting to the previous conclusion that men were more susceptible than women to SARS-CoV-2.[14, 15] This might be related to occupational exposures, for more men than women work as salesmen or market managers in seafood markets. As recorded, 66.0% of the patients in Huang’s report and 49% of the patients in Chen’s report had the history of exposure to the Huanan Seafood Market, and most of the affected patients were male workers.[13, 14] In contrast, no patient in our study had such exposure. All of these indicated a change of transmission mode outside Wuhan and that gender may not be a susceptible factor for COVID-19.

The median age of our patients was 46 years old, close to that of patients outside Wuhan as reported by Wu et al (46 years)[16]and Xu et al (41 years),[17] and younger than that of patients in Wuhan as reported by Wang et al (56 years)[18]and Chen et al (55 years).[14]Similarly, severe patients were much older than non-severe patients. This suggests that age may be an important risk factor for poor outcome. The role of age in COVID-19 seems to be similar to its role in SARS and MERS, which has been reported as an independent predictor of adverse outcome.[19, 20] T-cell and B-cell hypofunction and excessive production of type 2 cytokines in older people could lead to defect in inhibition of viral replication and stronger host innate responses with sustained cytokine storm, potentially leading to poor outcome.[21] Therefore, compromised immune function might be the major cause of higher mortality in older people infected by coronaviruses.

The proportion of severe cases in Shaanxi was close to that in Wuhan as reported by Wang et al.,[18] while the incidence of complications and mortality were considerably lower among Shaanxi patients than among the initially infected Wuhan patients.[13, 14, 18] Only two cases in our cohort needed mechanical ventilation. This might indicate that patients outside Wuhan had a much better prognosis than the first generation patients in Wuhan. What’s more, of the cases in Wuhan, those initially identified had a higher mortaility than those confirmed and treated later (15%[13] vs. 11%[14] vs. 4.3%[18]). This phenomenon was similar to that during the transmission of MERS-CoV, in which the global mortality of the first-generation MERS-CoV was about 35.5%, while that of the second-generation was around 20%.[22] Furthermore, the median interval from symptom onset to hospital admission in Shaanxi cases was shorter than in Wuhan cases (4.5 vs. 7 days).[13, 18] and the Shaanxi patients were younger than those in Wuhan (46 vs. 55-62 years).[14, 15, 18] These may be reasons for the notable reduction in mortality in Shaanxi cases.

The percentage of cases having fever in our cohort was lower than that reported in Wuhan.[13, 14, 18] In this regard, patients with normal temperature may be missed if the surveillance case definition focused heavily on fever detection. Compared with non-severe patients, severe patients more commonly had symptoms and signs such as cough, sputum, chest stuffiness, dyspnea, temperature above 38°C, respiratory rate above 21 breaths per minute, and heart rate above 100 beats per minute. The onset of symptoms and signs may assist physicians in identifying patients with greater severity.

Based on the radiological data, the incidences of bilateral pneumonia and pleural effusion were higher in severe cases than in non-severe cases, which suggested greater disease severity. Similar to what was reported by Sun et al,[23] in 54.7% (69/126) of the pneumonia cases, pulmonary fibrosis was found in later chest CT images when shadowing had been resolved, and the phenomenon was more common in severe patients than in non-severe patients. These findings consistently suggest that pulmonary fibrosis can be one of the sequelaes of COVID-19. It is necessary and important to explore how to prevent and reduce the occurrence of pulmonary fibrosis and how to manage the situation whenever it occurs in the treatment of COVID-19.

In terms of laboratory tests, different from cases in Wuhan, most Shaanxi patients had lymphocytes within the normal range, and only 38.1% showed lymphopenia. The lymphocyte absolute count in our cohort of patients (1.1×10^9^/L) was higher than that reported in Wuhan patients (0.6-0.8×10^9^/L).[13, 18, 24] This may be another reason for the lower mortality of Shaanxi cases as compared with of Wuhan cases. In severe cases, the lymphocyte count was lower and the incidence of lymphopenia was higher. These findings suggest that SARS-CoV-2 might mainly act on lymphocytes, especially T lymphocytes, and the severity of lymphopenia might reflect the severity of the disease. Furthermore, levels of inflammatoryparameters, such as CRP and ESR elevated in COVID-19 patients and were even higher in severe patients. The changes of these laboratory parameters illustrated that the virus invaded through respiratory mucosa and spread in the body, triggering a series of immune responses and inducing severe inflammation and cytokine storm in vivo.[25]

Few patients in our study had abnormal levels of alanine aminotransferase (ALT), aspartate aminotransferase (AST), and indirect bilirubin (IDBIL). The median level of DBIL in the patients elevated, and was even higher in severe patients. As reported, the potential mechanism of liver dysfunction in COVID-19 could be that the virus might directly bind to ACE2 positive bile duct cells.[26] Therefore, the liver abnormality of COVID-19 patients may not be caused by liver cell damage, but by bile duct cell dysfunction.

In addition, elevated glucose, LDH, HBDH, and pro-brain natriuretic peptide, as well as declined albumin, PaO_2_, and PaO_2_:FiO_2_, were more commonly seen in severe cases, suggesting greater disease severity.

The dynamic changes of six laboratory markers showed that baseline lymphocyte count was significantly higher in survivors than in the non-survivor patient. The lymphocyte count showed a gradual increase as the condition improved, but declined sharply when death occurred. Conversely, the IL-6 level displayed a downtrend in survivors, but continually rose to a very high level in the non-survivor patient. Hence, we assume that T cellular immune function might relate to mortality, and lymphocyte and IL-6 should be used as indicators for prognosis. Additionally, CRP, LDH, HBDH, and DBIL levels decreased as the condition improved in recovered patients, but increased rapidly as the condition worsened in the non-survivor patient and the critical case. These may be related to cytokine storm and bile duct cell dysfunction induced by virus invasion.

Most patients (96.3%) in our study received antiviral therapy, including lopinavir/ritonavir (64.9%), interferon alpha inhalation (50.7%), arbidol (42.5%), ribaviron (32.8%) and chloroquine (2%).Up to now, no specific treatment has been recommended for COVID-19 infection except for optimal supportive care. A previous study showed that combination of lopinavir and ritonavir was associated with substantial clinical benefit for SARS infection.[27] Another study claimed that remdesivir had a good therapeutic effect on COVID-19.[28] Currently, randomised clinical trials for lopinavir/ritonavir (ChiCTR2000029308) and intravenous remdesivir (NCT04257656, NCT04252664) in treatment of COVID-19 are in progress.[24] Meanwhile, COVID-19 vaccine is highly expected. Ongoing efforts are needed to explore effective therapies for this emerging acute respiratory infection.

### Limitations of study

This study has some limitations. First, as only COVID patients in Shaanxi were recruited, our conclusions need to be further verified by recruiting larger number of cases of other provinces or cities, outside Wuhan. Second, there were 2 critically ill cases with 1 non-survivor in our study, thus dynamic observations of laboratory parameters between non-survivor and survivor, recovered cases and critically ill cases were just descriptive analysis. Larger number of critically ill cases are needed to verify our observation.

## Conclusion

In summary, the present study identified that the imported and second-generation COVID-19 cases in Shaanxi had a better prognosis in comparation with initial or first-generation cases in Wuhan, with less complications, lower fatality, and higher discharge rate. These differences may be related to the shorter interval from symptom onset to hospital admission, younger age, and higher lymphocyte count of patients in Shaanxi. Lymphocyte count and IL-6 level could be used as indicators for evaluating prognosis. CRP, LDH, HBDH, and DBIL levels could help estimate the severity and development tendency of the disease. Pulmonary fibrosis was found in later chest CT images in more than half of the pneumonia cases and should be taken into account.

## Data Availability

No additional data available.

## Notes

### Contributors

PYS, GXR, JY, ZQL, SJD, ML, SSW, XFX, FPC, YJL, CYL, XHY contributed equally to this paper. Concept and design: PYS, GXR, JY, MWC; Acquisition, analysis, or interpretation of data: PYS, GXR, JY, ZQL, SJD, ML, SSW, XFX, FPC, YJL, CYL, XHY; Drafting of the manuscript: PYS, GXR, JY, ZQL, SJD, ML, SSW, XFX, FPC, YJL, CYL, XHY, MWC; Critical revision of the manuscript for important intellectual content: PYS, MWC; Statistical analysis: PYS, ZFX, ZXW; Administrative, technical, or material support: PYS, ZFX, ZXW; Supervision: PYS, MWC.

### Ethical approval

This study was approved by the Ethics Committee of the First Affiliated Hospital of Xi’an Jiaotong University (XJTU1AF2018LSK-021).

## Acknowledgments

We are grateful to all health-care workers involved in the diagnosis and treatment of COVID patients. We thank all our colleagues who helped us during the current study.

